# Respiratory Failure in Covid19 is associated with increased monocyte expression of complement receptor 3

**DOI:** 10.1101/2020.05.31.20118638

**Authors:** Rajeev Gupta, Vanya Alasdair, Bryan Williams Gant, Tariq Enver

## Abstract

A key question in COVID-19 infection is why some previously healthy patients develop severe pulmonary failure and some ultimately die. Initial pulmonary failure does not exhibit classical features of ARDS; hypercoagulability is a common laboratory feature, and pulmonary thrombotic microangiopathy has been reported post mortem1,2,3. Biomarkers cannot robustly identify such patients pre-emptively and no specific interventions exist to mitigate clinical deterioration. Mononuclear phagocytic cells are key immune cells and bind fibrinogen through the CD11b/CD18 dimer CR3, whose activated form can initiate microthrombus formation. Accordingly, we profiled circulating monocyte CD11b/CD18 cell surface density from COVID-19 infected adults who were (i) symptomatic but breathless, (ii) requiring ventilatory support, and (iii) recovering following ICU care for hypoxia.

---

A key question in COVID-19 infection is why some previously healthy patients develop severe pulmonary failure and some ultimately die. Initial pulmonary failure does not exhibit classical features of ARDS; hypercoagulability is a common laboratory finding, and pulmonary thrombotic microangiopathy has been reported *post mortem*^1,2,3^. Biomarkers cannot robustly identify such patients pre-emptively and no specific interventions exist to mitigate clinical deterioration. Mononuclear phagocytic cells are key immune cells and bind fibrinogen through the CD11b/CD18 dimer CR3, whose activated form can initiate microthrombus formation. Accordingly, we profiled circulating monocyte CD11b/CD18 cell surface density from COVID-19 infected adults who were (i) symptomatic but breathless, (ii) requiring ventilatory support, and (iii) recovering following ICU care for hypoxia.

## Methods

We repurposed a multiparameter flow cytometry assay, used routinely in our institution for the diagnosis of leukaemia, to measure the cell surface expression density of CD11b and CD18 in peripheral blood monocytes from three clinically distinct groups of COVID19 patients (29 in total), and compared them to non-COVID-19 patients requiring ventilatory support, with the historical group of healthy controls used for initial test calibration as reference. Analysis was performed using surplus material from routine CBC specimens. Sample preparation was with the TQ-Prep whole blood lysis system (Beckman Coulter), and antibody staining was with a Duraclone lyophilised antibody panel (B38683, Beckman Coulter, Table 1). A minimum of 50000 events were collected: monocytes were defined as CD45^+^CD13^+^CD33^++^CD15^+/−^SSc^int^ cells. The expression level of each antigen was calculated as its corrected Mean Fluorescence Intensity (MFI): [raw value arithmetic mean fluorescence in monocytes – raw value arithmetic mean fluorescence in CD45^++^CD2^+^CD56- SSc^lo^ T cells]. We defined “hypoxic” patients as requiring ICU Level 3 ventilatory support within 15 days of a PCR diagnosis of SARS-CoV2 infection; “convalescing” as having recovered following ICU care for COVID with minimal or absent oxygen requirements, but at least 30 days from initial diagnosis; and “well” as having mild symptoms prompting a positive PCR test within the previous 10 days, but who were neither ever breathless nor hospitalised. No patient in this study had a prior history of malignancy or immunosuppression.

**Table 1.** Antigen specificities with respective clone identities and fluorochromes in the Duraclone B38683 antibody panel

| Antigen | CD2 | CD11b | CD13 | CD15 | CD18 | CD33 | CD34 | CD45 | CD56 | CD117 |
| --- | --- | --- | --- | --- | --- | --- | --- | --- | --- | --- |
| Clone | 39C1.5 | Bear1 | Immu103.44 | 80H5 | 7E4 | D3HL60.251 | 581 | J33 | N901 | 104D2D1 |
| Fluorochrome | APC-a750 | FITC | PC5.5 | PB | PE | PC7 | ECD | KrO | APC-A700 | APC |

## Results

Table 2 shows clinical and haematological parameters and Figure 1 the levels of monocyte CD11b and CD18 for each COVID-19 group and the two non-COVID-19 cohorts. CBC parameters were not significantly different, however expression of both CD11b and CD18 was higher in COVID-19 patients who were hypoxic requiring respiratory support. Convalescing patients had intermediate levels of CD11b, but CD18 had returned to normal. Levels were not elevated in patients who had SARSCoV2 infection but remained well, and those requiring ventilatory support for reasons other than COVID-19. Consistent with others, we found that hypoxic COVID-19 patients had lymphopenia, but this was also seen in non-COVID hypoxic patients, and likely represents a stress response.

**Table 2.** Laboratory parameters of patient groups. COVID “hypoxic”, COVID “convalescing” and non-COVID “hypoxic” cohorts had significantly lower CD3^+^4^+^ and CD3^+^8^+^ T cell counts, and significantly higher serum LDH levels than the COVID “well” group (*p*<*0.05*). The difference in D-Dimer between the three COVID groups was not significant.

| Parameter<br>(normal range) |  | COVID<br>"well"<br>n=11 | COVID<br>"hypoxic"<br>n=10 | COVID<br>"convalescing"<br>n=8 | Non-COVID<br>"hypoxic"<br>n=5 |
| --- | --- | --- | --- | --- | --- |
| Total WCC<br>(3.0-10.0 x10<sup>9</sup>/L) | mean<br>range | 7.01<br>(3.73-10.36 x10<sup>9</sup>/L) | 7.00<br>(2.01-13.1 x10<sup>9</sup>/L) | 8.54<br>(3.01-14.31 x10<sup>9</sup>/L) | 12.12<br>(3.48-34.60 x10<sup>9</sup>/L) |
| Monocytes<br>(0.1-1.0 x10<sup>9</sup>/L) | mean<br>range | 0.54<br>(0.32-0.84 x10<sup>9</sup>/L) | 0.35<br>(0.13-0.63 x10<sup>9</sup>/L) | 0.54<br>(0.17-1.20x10<sup>9</sup>/L) | 0.56<br>(0.26-1.00 x10<sup>9</sup>/L) |
| CD3<sup>+</sup>4<sup>+</sup> T cells<br>(0.48-1.64 x10<sup>9</sup>/L) | mean<br>range | 0.80<br>(0.43-1.51 x10<sup>9</sup>/L) | 0.28<br>(0.11-0.41 x10<sup>9</sup>/L) | 0.74<br>(0-35-1.87 x10<sup>9</sup>/L) | 0.11<br>(0.01-0.42 x10<sup>9</sup>/L) |
| CD3<sup>+</sup>8<sup>+</sup> T cells<br>(0.21-1.09 x10<sup>9</sup>/L) | mean<br>range | 0.46<br>(0.18-0.95 x10<sup>9</sup>/L) | 0.10<br>(0.02-0.22 x10<sup>9</sup>/L) | 0.18<br>(0.00-0.33 x10<sup>9</sup>/L) | 0.24<br>(0.01-0.97 x10<sup>9</sup>/L) |
| LDH<br>(135-226 IU/L) | mean<br>range | 205<br>(149-254 IU/L) | 400<br>(256-561 IU/L) | 380<br>(255-563 IU/L) | 312<br>(199-576 IU/L) |
| D-Dimer<br>(0-550 µg/L) | mean<br>range | 887<br>(190-8260 µg/L) | 4128<br>(810-17070 µg/L) | 2524<br>(540-8330 µg/L) | not available |

**Figure 1.**
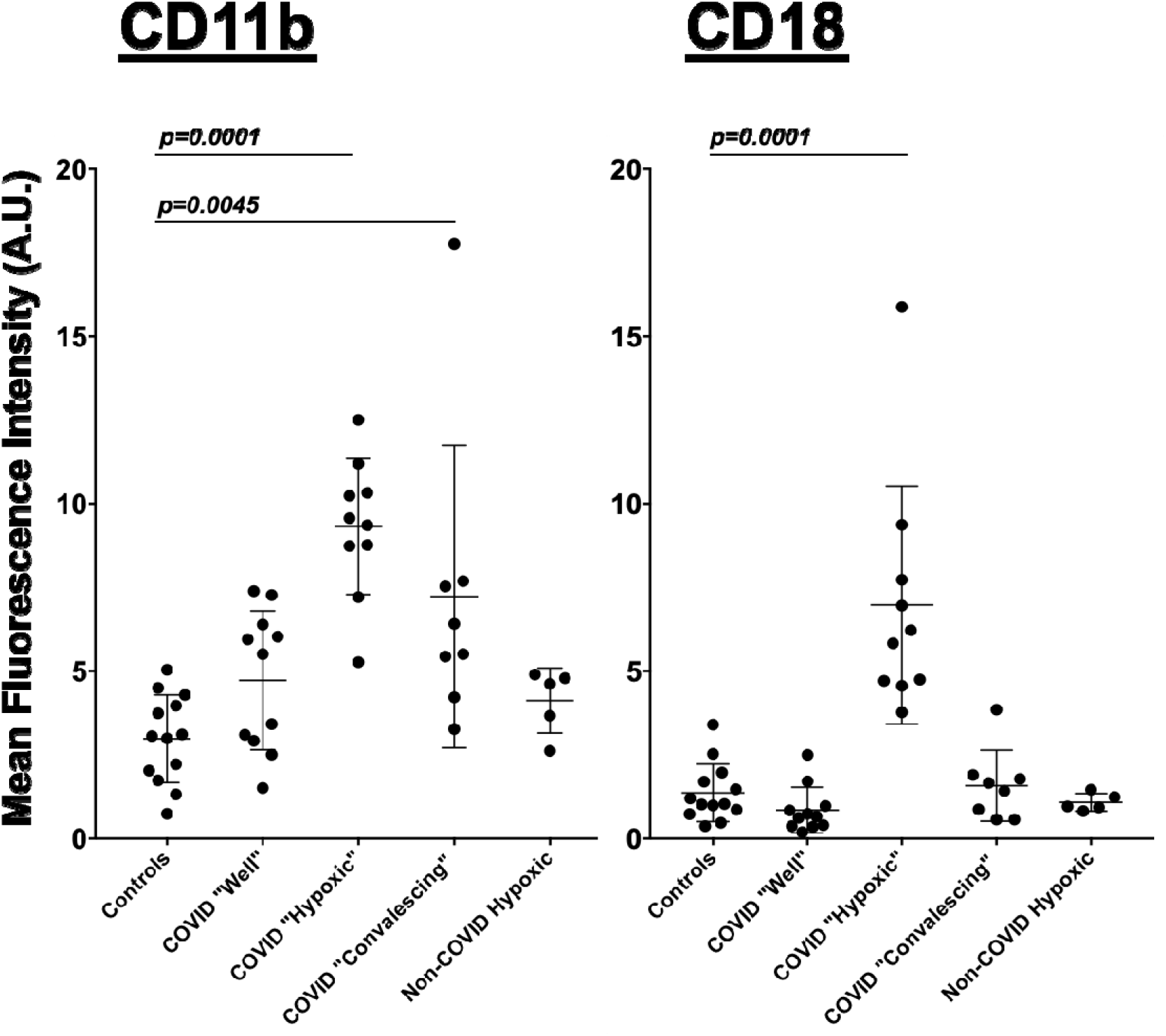
Mean Fluorescence intensity of CD11b and CD18 expression (Arbitrary Units) in peripheral blood monocytes. Controls are the historical cohort of healthy volunteers with normal blood counts used for validation of the assay. Horizontal bars indicate groups with significantly different expression levels and relevant p values.

## Discussion

Monocytes mediate initial immune responses to pathogens, induce inflammation following tissue damage and regulate haemostasis at sites of injury^4^. CD11b and CD18, which together constitute Complement Receptor 3 (CR3) are key to this^5^. We demonstrate that monocyte CR3 is significantly increased in COVID-19 patients with hypoxia compared with those without respiratory symptoms and non-COVID hypoxic patients. This hitherto unappreciated observation tables monocyte activation as a biomarker for respiratory complications in COVID-19, possibly by driving thrombotic microangiopathy due to increased binding of fibrinogen. Although adaptive immune responses to SARS-CoV2 determine subsequent immunity, innate immune responses dictate the early clinical course, and our findings encourage serial tracking of monocyte CR3 expression in individual patients with COVID-19. CR3 may be targeted therapeutically; both non-specifically by early anticoagulation, or directly using several licensed drugs^5^. Such approaches may reduce the likelihood of respiratory failure and could inform a preventative approach to the management of COVID19- mediated respiratory failure in the future.

## Data Availability

All data is provided in the manuscript

## Author Contributions

RG and VAG had full access to all the data in the study and take responsibility for the integrity of the data and the accuracy of the data analysis.

### Concept and design

RG, VAG, TE

### Acquisition, analysis, or interpretation of data

All authors

### Drafting of the manuscript

RG, VAG

### Critical revision of the manuscript for important intellectual content

All authors

### Administrative, technical, or material support

All authors

### Supervision

All authors

### Funding/Support

The study was funded by UCL Biomedical Research Centre’s emergencyCOVID-19 funds and Blood Cancer UK. We are also extremely grateful to Sir Peter Woodfor his philanthropic donation towards essential Research laboratory equipment.

## References

1. Richardson S, Hirsch JS, Narasimhan M, et al. Presenting Characteristics, Comorbidities, and Outcomes Among 5700 Patients Hospitalized With COVID-19 in the New York City Area JAMA. Published online April 22, 2020. doi:10.1001/jama.2020.6775

2. Varga Z, Flammer AJ, Steiger P, et al Endothelial cell infection and endotheliitis in COVID-19. Lancet Published online April 17, 2020 https://doi.org/10.1016/S0140-6736(20)30937-5

3. Magro C,,Mulvery JJ, Berlin D, et al. Complement associated microvascular injury and thrombosis in the pathogenesis of severe COVID-19 infection: A report of five cases, Translational Research (2020), doi: https://doi.org/10.1016/j.trsl.2020.04.007

4. Class CK and Natoli G. Molecular control of activation and priming in macrophages. Nature Immunology 2016; 17:26–33

5. Vorup-Jensen T and Jensen RK. Structural Immunology of Complement Receptors 3 and 4. Front. Immunol. 2018; 9:2716.

